# Wastewater-based surveillance of respiratory syncytial virus epidemic at the national level in Finland

**DOI:** 10.1101/2023.09.04.23295011

**Authors:** Annika Länsivaara, Kirsi-Maarit Lehto, Rafiqul Hyder, Oskari Luomala, Anssi Lipponen, Anna-Maria Hokajärvi, Annamari Heikinheimo, Tarja Pitkänen, Sami Oikarinen, WastPan Study Group

## Abstract

The purpose of this study was to evaluate the potential of wastewater-based surveillance in the monitoring of epidemics at the national level in Finland. To discover the correlation of wastewater data and register data, the 2021–2022 respiratory syncytial virus (RSV) epidemic in Finland was analyzed from wastewater and the Finnish National Infectious Diseases Register. The study was performed using samples that were collected monthly from May 2021 to July 2022 from ten wastewater treatment plants that cover 40% of the Finnish population. Respiratory syncytial virus detection in 24-h composite samples of influent wastewater was performed using RT-qPCR. Respiratory syncytial virus wastewater data were positively correlated with the National Infectious Diseases Register data for the sampling week (correlation coefficient, CC min = 0.412, max = 0.865). Furthermore, the cumulative incidence of respiratory syncytial virus from the sampling week to three weeks afterward was strongly correlated with the wastewater data (CC min = 0.482, max = 0.814), showing the potential of wastewater-based surveillance for use in estimating the course of the epidemic. When the register-based incidence of RSV was at least four cases/100,000 persons/week in the sampling week, it was detected in all wastewater samples. This study showed that wastewater surveillance is useful in the surveillance of respiratory syncytial virus epidemics, and its potential in the surveillance of other epidemics should be explored further.

**Synopsis:** Environmental surveillance has been previously used to monitor viruses such as poliovirus and SARS-CoV-2. This study shows that wastewater-based surveillance can be used to surveil the respiratory syncytial virus and its epidemics, at a national level.

## Introduction

Respiratory syncytial virus (RSV) is a single-stranded enveloped RNA virus that belongs to the paramyxoviruses (Chen et al., 2021). It includes A and B subtypes (Laham et al., 2017). Typically, RSV causes lower respiratory tract infections for all ages, while premature infants, young children, immunocompromised individuals, and the elderly are at a higher risk (Kaler et al., 2023). RSV can lead to pneumonia or bronchiolitis (Jain et al., 2023). In fact, in children under 2 years old, RSV is the most common cause of viral pneumonia and acute bronchiolitis (Perk and Özdil, 2018). RSV has been reported to place an even greater burden on children’s healthcare than COVID-19, and during the RSV epidemic in Finland in 2022, it was the most common cause of hospital admissions in children (Helsinki University Hospital, 2022). In low-and middle-income countries, RSV is one of the leading causes of mortality and morbidity in children up to five years old (Staadegaard et al., 2021). In that age group, it is estimated that RSV causes 3.2 million hospital admissions and 60,000 in-hospital deaths yearly (Andeweg et al., 2021). Thus far, there is no effective treatment or prevention for the diseases caused by RSV (Perk and Özdil, 2018). The infection can also be contracted more than once (Glezen et al., 1986). RSV spreads as a droplet infection (Bardsley et al., 2023). Its incubation period is typically from four to six days (Jain et al., 2023). Based on an earlier study, the virus was found to be shed for a median of 11.5 days, and more than 75% of family members of RSV-infected children were shown to have an upper-respiratory-tract infection during follow-up (von Linstow et al., 2006).

RSV has been found in the stool, nasal secretions, saliva, and sweat of infected persons (von Linstow et al., 2006). Multiple studies have reported that another respiratory virus, SARS-CoV-2, can be detected in stool even during the recovery period, with mild symptoms (Wölfel et al., 2020). These findings suggest that respiratory viruses can be detected in the secretions of even asymptomatic patients. Thus, wastewater-based surveillance (WBS) could be used to monitor RSV infections in the population. In contrast to clinical surveillance, which only detects cases that seek treatment, WBS could reveal the community-level disease prevalence, including subclinical infections (Robins et al., 2022). For example, based on wastewater data, the spread of COVID-19 has been shown to be much higher than it was according to clinical data (Wu et al., 2020). Respiratory viruses, such as RSV, may cause mild symptoms, and these cases are not clinically tested. Furthermore, many respiratory viruses cause similar symptoms, and therefore, without laboratory confirmation, RSV may be diagnosed incorrectly. Thus, the RSV epidemic is heavily underreported in clinical testing. WBS represents the entire population of the wastewater treatment plant (WWTP) catchment area, and it shows the unbiased virus load in the population.

WBS was implemented in the surveillance of viruses even before the COVID-19 pandemic, as poliovirus has been monitored in wastewater as a part of poliovirus eradication using environmental surveillance (Hovi et al., 2012). WBS has been used as an early-warnings tool for epidemics and outbreaks. SARS-CoV-2 RNA detected in wastewater was shown to predict upcoming COVID-19 cases by five days and hospitalization due to COVID-19 by eight days (Galani et al., 2022). During a norovirus epidemic in Sweden, the largest amount of the virus was detected in wastewater two weeks before the highest number of clinical cases (Hellmér et al., 2014). Furthermore, for influenza, the lead time from wastewater detection to community outbreak was shown to be 17 days (Mercier et al., 2022).

This is the first study to report the testing and detection of RSV in municipal wastewater in Finland. In this study, we determined the RNA copy numbers of RSV in wastewater influent using reverse-transcription quantitative PCR (RT-qPCR). The wastewater samples used for the study were collected monthly between May 2021 and July 2022 from ten wastewater treatment plants (WWTPs) in Finland, which cover 40% of the Finnish population. This study allows the evaluation of the temporal and spatial changes of the RSV epidemic in Finland, and it can be used to evaluate the potential of WBS in the surveillance of epidemics at the national level.

## Materials and Methods

### 2.1 Wastewater sampling

This study was performed as part of the WastPan consortium, which consists of Tampere University, the Finnish Institute for Health and Welfare (THL), and the University of Helsinki, from 2020 to 2023 (Lehto et al., 2023). The WastPan consortium studies wastewater-based surveillance and antimicrobial resistance genes and is developing tools for their detection. This national RSV surveillance study includes ten WWTPs located in Espoo (Suomenoja WWTP, 390,000 inhabitants), Helsinki (Viikinmäki WWTP, 860,000 inhabitants), Kuopio (Lehtoniemi WWTP, 91,000 inhabitants), Lappeenranta (Toikansuo WWTP, 63,000 inhabitants), Oulu (Taskila WWTP, 200,000 inhabitants), Pietarsaari (Alheda WWTP, 31,000 inhabitants), Rovaniemi (Alakorkalo WWTP, 55,000 inhabitants), Seinäjoki (Seinäjoen keskuspuhdistamo WWTP, 55,000 inhabitants), Tampere (Viinikanlahti WWTP, 200,000 inhabitants), and Turku (Kakolanmäki WWTP, 300,000 inhabitants). These WWTPs cover 40% of the Finnish population, enabling national-level surveillance. Samples were collected from these WWTPs monthly from May 2021 to July 2022. Influent wastewater was collected for 24 h as a composite sample, after which a 1-liter fraction of the sample was shipped in cool boxes. The samples arrived at the laboratory within 24–48 h of sampling.

### 2.2 Concentration of wastewater samples

The concentration of the samples was performed immediately after the samples arrived at the laboratory. An aliquot of 100 ml was taken from the 1 L sample. Then, the aliquot was concentrated as in Hokajärvi et al. (2021). First, particles that could block an ultrafilter were removed by centrifuging the samples at 4,654 g for 30 min without a brake. Afterward, the supernatant was concentrated with a Centricon® Plus-70 centrifugal ultrafilter with a cut-off of 100 kDa (Millipore, Cork, Ireland) by centrifuging the samples at 3,500 g for 15 min. The concentrate was collected by centrifuging the filter at 1,000 g for 2 min.

### 2.3 RNA extraction

Immediately after the samples were concentrated, RNA was extracted from 300 µL of the concentrates. The RNA extraction was performed using PerkinElmer Chemagic Viral DNA/RNA 300 (Wallac Oy, Turku, Finland) RNA extraction kit according to the manufacturer’s protocol. The kit used an automated extraction process based on the magnetic-bead isolation of RNA. The elution volume was 100 µl.

### 2.4 RT-qPCR detection of RSV

RSV specific RT-qPCR was performed using TaqMan Fast Virus 1-Step Mastermix (TaqMan RT-qPCR; Applied Biosystems by Thermo Fisher Scientific, Vilnius, Lithuania). The RT-qPCR was performed according to the manufacturer’s recommendations, using a total volume of 25 µL. The reaction mixture included 6.25 µL of the TaqMan Fast Virus 1-step mastermix, a 900 nM forward primer, a 900 nM reverse primer, a 300 nM probe, and 5 µL of template. The sequences of the primers and probes were 5’-CTCCAGAATAYAGGCATGAYTCTCC-3′ for the forward primer, 5’-GCYCTCCTAATYACWGCTGTAAGAC-3′ for the reverse primer, and 5’-6FAM/GTAATAACCAAATTAGCAGCAGGRGATAGA/TAM-3’ for the probe. The primers and probes were modified from Salimi et al. (2021) and targeted both the A and B subtypes of RSV. The cycling conditions for the RT-qPCR reactions were as follows: reverse transcription at 50 ⁰C for 5 min, one cycle of RT-inactivation and initial denaturation at 95 ⁰C for 20 s, 50 cycles of denaturation at 95 ⁰C for 15 s, and annealing and extension with fluorescence data collection at 58 ⁰C for 1 min. The samples were run as duplicates. A negative control for RNA extraction and negative and positive controls for RT-qPCR were run on each plate.

The quantification of the RSV gene copy numbers was performed using a dilution series of an RSV control from 10² to 10⁶. The RSV control was prepared *in vitro* in the pET3A plasmid using biomolecular technology. Initially, the primer sequences of RSVB were commercially cloned into the pET3A plasmid using GenScript (The Netherlands). Later, the recombinant DNA plasmid was transformed into competent DH5 alpha E. coli cells. The transformed cells were screened on agar media. Selected clones were further grown in liquid medium, and the plasmids were isolated using the commercial GeneJet plasmid Midi Prep kit (ThermoFisher Scientific, Lithuania) and sequenced by Macrogen (The Netherlands). Plasmids with correct sequences were digested with the Cla I restriction enzyme (Promega, USA) at the 3′-end of the DNA before transcription with MegaScript T7 (Invitrogen by ThermoFisher Scientific, Lithuania). RNA transcripts were purified from the untranscribed DNA using an RNeasy Minikit (Qiagen, Germany). The purified RNA was quantified using a Nanodrop ND-1000 UV-Vis spectrophotometer (ThermoFisher Scientific), and a series of serial dilutions (2.07*10^2–10^6) in RNA storage solution (ThermoFisher Scientific; 1 mM sodium citrate, pH 6.4) and RNasin (ThermoFisher Scientific, Lithuania; 20 U/μL) were prepared. The dilutions were aliquoted and stored at −80°C. (Lehto et al., 2023). To continue, the 95% limit of detection (LOD) was calculated for the RT-qPCR analysis using regression probit analysis (Stokdyk et al., 2016). The LOD was determined to be 13.6 copies/µL.

### 2.5 Normalization of the RSV wastewater data

The RSV gene copy (GC) number in the wastewater was normalized to the flow and population of each WWTP via a method described previously (Tiwari et al., 2022). The RSV copies per m³ were multiplied with 24 h of inflow in m³ for a WWTP. The result was divided by the estimated population served by the WWTP. The final normalized value for each sample was reported in gene copies/1,000 persons/24 h.

### 2.6 Laboratory-confirmed RSV cases

Clinical RSV data were collected from the National Infectious Diseases Register (NIDR), which is managed by the Finnish Institute for Health and Welfare. The NIDR includes laboratory-confirmed RSV cases that have been reported by clinical laboratories. For each of the WWTPs, a 7-day incidence of RSV was calculated using the RSV cases in the WWTP catchment area. The number of RSV cases in the catchment area in a week was divided by the estimated number of inhabitants in the WWTP catchment area and then multiplied by 100,000 to obtain the RSV incidence in terms of RSV cases/100,000 persons/week. In addition, the cumulative incidence from the sampling week to 3 weeks afterward was calculated. The sum of RSV cases in the catchment area in the sampling week and 1, 2, and 3 weeks after sampling was divided by the estimated number of inhabitants in the WWTP catchment area and then multiplied by 100,000 to obtain the RSV cumulative incidence in terms of RSV cases/100,000 persons/4 weeks after sampling. The incidence and cumulative incidence were both correlated with the RSV gene copies in the wastewater.

### 2.7 Statistical analysis

All statistical analyses were calculated using IBM® SPSS® software, Version 28.0.1.0 (142). According to a Saphiro–Wilk test, the data were not normally distributed. Thus, Kendall rank-correlation coefficients were used to evaluate correlations between two nonparametric ranked samples, the incidence of RSV disease cases based on NIDR data, and the gene copies of RSV in wastewater.

## 3 Results and Discussion

### 3.1 Uniqueness of the study

This study presents the results of unique RSV surveillance using wastewater. To our knowledge, only five studies have been published on the surveillance of RSV in wastewater, which were limited to smaller geographical areas and not performed at the national level (Ahmed et al., 2023; Ando et al., 2023; Boehm et al., 2022; Hughes et al., 2022; Toribio-Avedillo et al., 2023). We present the results of national RSV surveillance. This study includes ten WWTPs from Finland. The WWTPs are geographically representative of Finland and their catchment area includes 40% of the Finnish population (Lehto et al. 2023). The population coverage of this study is similar to the coverage of Finnish national SARS-CoV-2 surveillance, which covers 44% of the population and in which nine WWTPs are monitored weekly and, in addition, two WWTPs are monitored monthly (Finnish Institute for Health and Welfare, 2023). This study was also temporally extensive, as samples were collected for 18 months, from May 2021 to July 2022. Thus, our data allow for the generalization of the results of this study, both spatially and temporally, to the whole of Finland.

Previous studies on the wastewater surveillance of RSV have used RSV cases detected in the city or the state of the WWTP. Our study is the first to match the incidence of RSV to the catchment areas of each WWTP, meaning that the study populations for the clinical data and wastewater data are the same. This allows a more accurate comparison of clinical data and wastewater data than using citywide or statewide incidences.

### 3.2 The incidence of RSV based on the Finnish National Infectious Diseases Register

The seasonality of RSV in Finland was evaluated based on NIDR data. Typically, in Finland, the RSV season begins in October or November, reaches a peak in February or March, and ends in April or May. The RSV epidemic in 2020 and 2021 did not follow the typical seasonality in Finland, most likely due to the COVID-19 restrictions. Between October 2020 and March 2021, only 26 cases of RSV were reported in the NIDR. The RSV season was also shorter in 2020– 2021 as compared to the typical seasonality of RSV in Finland (Kuitunen et al., 2020). This untypical seasonality for respiratory viruses other than SARS-CoV-2 was also noted for influenza. In the WHO European Region, the number of positive influenza specimens in the 2020–2021 influenza season was 99.8% lower than in any previous season (Adlhoch et al., 2021).

In Finland, the 2021–2022 RSV epidemic was noted to start in October, and the peak occurred in December; by April, the epidemic was over (Figure 1). The first reported cases of RSV during the follow-up period occurred in May in Helsinki, Kuopio, and Tampere; in June in Espoo and Seinäjoki; in July in Turku; in September in Oulu; in October in Rovaniemi; in November in Lappeenranta; and in December in Pietarsaari. In most of the cities, the incidence of RSV exceeded five cases/100,000 persons/week in November, while in Pietarsaari, Rovaniemi, and Tampere, this occurred in December. The peak of the epidemic occurred in December in most of the cities, while in Kuopio and Pietarsaari, it occurred in January. The incidence of RSV decreased to under five cases/100,000 persons/week in January in Espoo, Helsinki, Tampere, and Turku; in February in Lappeenranta and Oulu; in March in Kuopio and Pietarsaari; in April in Rovaniemi; and in May in Seinäjoki. The last reported case of the follow-up period was in May in Lappeenranta and Tampere, in June in Rovaniemi, and in July in Kuopio. In the other cities, a few cases were still being reported at the end of the follow-up period.

**Figure 1.**
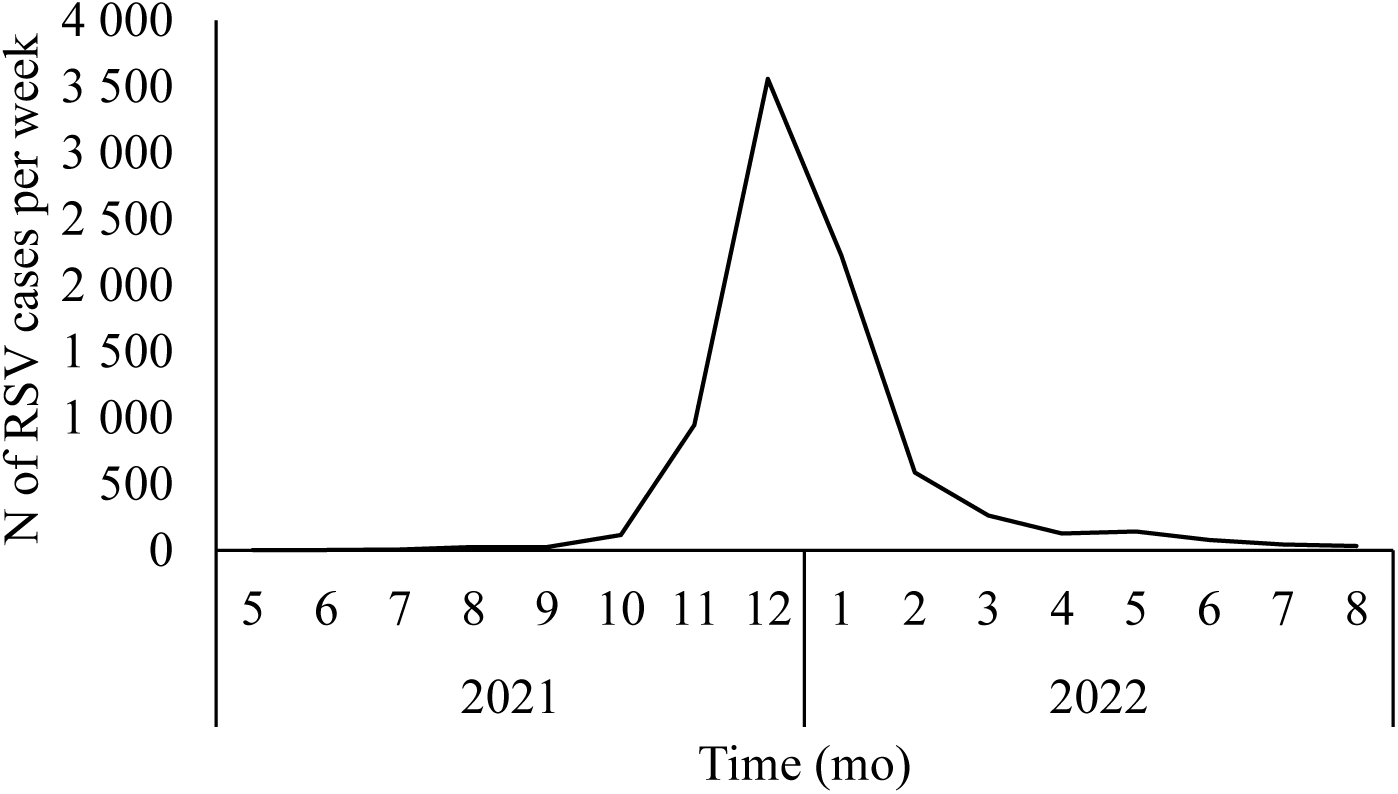
Laboratory confirmed RSV cases in Finland during the 2021-2022 epidemic according to the national infectious diseases register.

According to the NIDR data, the RSV epidemic began and ended, and the peak occurred at different timepoints in different WWTP catchment areas. Furthermore, the duration of the epidemic differed between cities. This highlights the importance of using clinical data that are matched to WWTP catchment areas and not statewide or nationwide clinical cases. Using clinical data that are matched to WWTP catchment areas allows for a more accurate comparison of clinical-and wastewater data.

### 3.3 RSV in wastewater

Of the 150 wastewater samples tested, 36% (N = 55) were positive for RSV. A positive relationship was seen between RSV incidence and RSV detected in wastewater samples. When RSV cases were reported in the population during the wastewater-sample collection week, RSV was detected in 70% of wastewater samples. However, 14% (N = 10) of the wastewater samples were positive for RSV when no RSV cases were reported in the WWTP area. Eight of these samples were collected 1–2 weeks prior to RSV cases being reported in the WWTP area. This shows the capability of wastewater surveillance to predict RSV cases and monitor epidemics during periods of low incidence. It also reflects cases without symptoms or with only mild symptoms that are not tested.

RSV was detected in all samples when the incidence of RSV exceeded four cases per 100,000 persons in the sampling week (Table 1). A similar sensitivity was observed for influenza A in Finland. When the incidence of influenza A exceeded five cases per 100,000 individuals in the sampling week, the virus was detected in wastewater samples (Lehto et al. 2023). These results suggest that wastewater surveillance is a sensitive method when used for the surveillance of viruses.

**Table 1.** A positive association between the RSV incidence (RSV cases per 100 000 persons per week) and the amount of RSV positive wastewater samples was noted.

| RSV incidence per 100<br>000 | Detected in wastewater<br>(%) | N of positive samples/N of<br>samples |
| --- | --- | --- |
| 0 | 14 | 10/74 |
| >0-2 | 32 | 12/38 |
| 2-4 | 64 | 9/14 |
| >4 | 100 | 24/24 |

The peak of the RSV epidemic in both wastewater and NIDR data followed a similar pattern (Figure 2). The first RSV-positive wastewater sample was detected in August in Espoo and Tampere; in September in Oulu and Rovaniemi; in October in Seinäjoki; in November in Helsinki, Lappeenranta, and Turku; and in December in Kuopio and Pietarsaari. In all the cities, cases were reported in the NIDR before wastewater detection. Still, RSV could be detected in wastewater before the incidence exceeded five cases/100,000 persons/week in Espoo, Oulu, Rovaniemi, and Tampere. Furthermore, in Helsinki, Lappeenranta, Pietarsaari, and Turku, wastewater was found to be RSV positive in the same month that the incidence of RSV exceeded 5/100,000/week. The epidemic could be detected in wastewater before its peak (NIDR data) for all WWTPs. In all cities, wastewater samples were still found to be positive after the incidence of RSV fell below 5/100,000/week permanently. The peak of the epidemic occurred at different timepoints according to the wastewater and NIDR data. The wastewater data show the peak of the epidemic to be in January in eight of the cities, while in Helsinki, the peak occurred in December. In Rovaniemi, it occurred in February. According to the NIDR data, the peak was earlier in most cities and at the same time in Kuopio and Pietarsaari. The last positive RSV wastewater sample from the follow-up period occurred in February in Espoo, Helsinki, and Lappeenranta; in March in Tampere; in April in Rovaniemi; in June in Kuopio, Oulu, Seinäjoki, and Turku; and in July in Pietarsaari. However, it needs to be taken into consideration that wastewater was collected only monthly, and the sampling interval was from 3 to 6 weeks. This did not allow for the exact timing of the initiation, peak, and end of the epidemic according to the wastewater data. More frequent wastewater sampling would increase the accuracy of the wastewater data and its temporal association with the incidence data for RSV cases.

**Figure 2.**
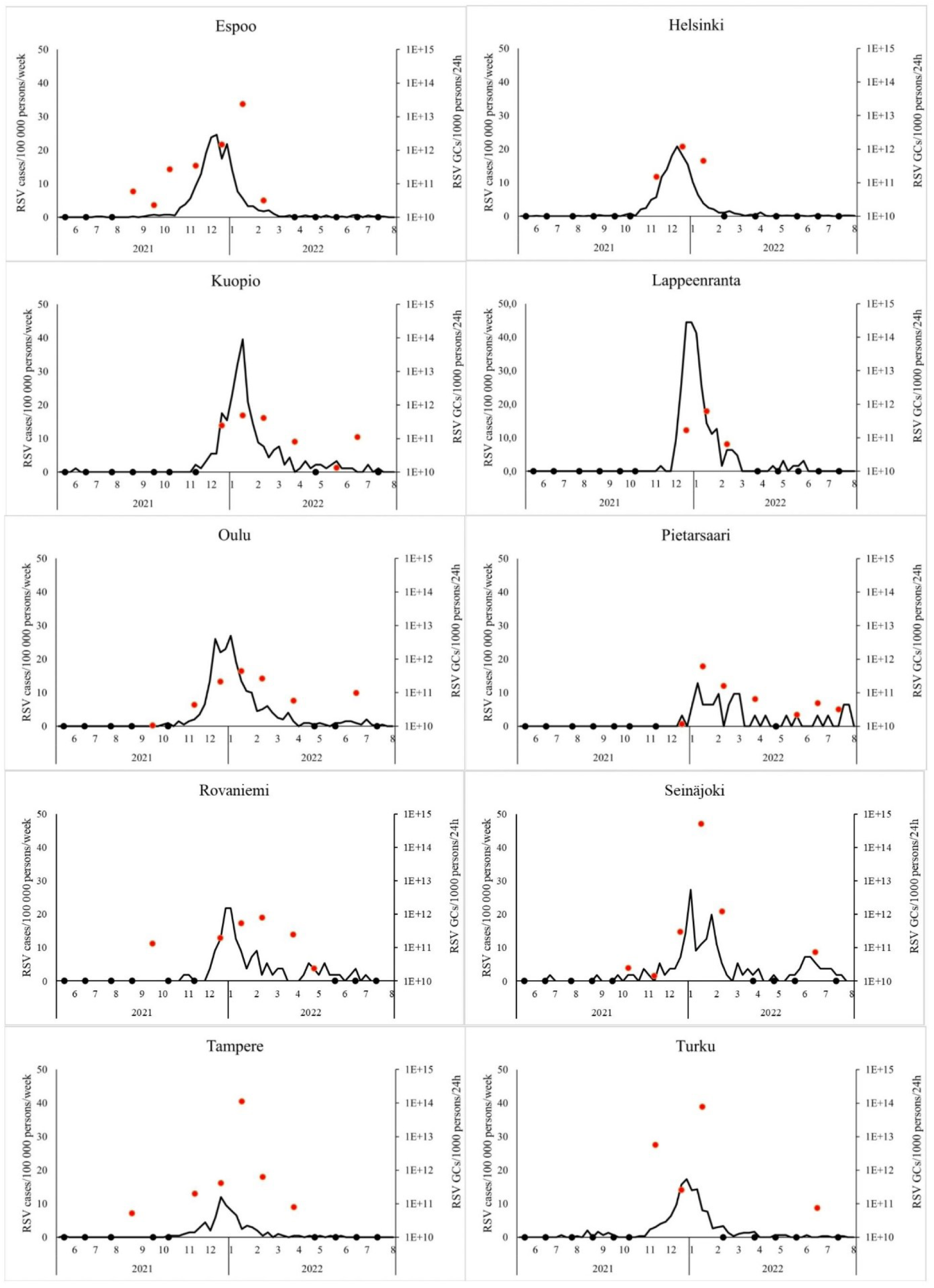
The positive association between the incidence of RSV (number of RSV cases per 100 000 persons per week) and RSV detected in wastewater (RSV gene copies per 1000 persons per 24h). The black line shows the incidence of RSV according to NIDR across the time series of the study, the red dots represent wastewater samples that were RSV positive, and the black dots represent wastewater samples that were RSV negative.

### 3.4 Correlation with the NIDR data

The RSV wastewater data were shown to have a positive correlation with the NIDR RSV data (Table 2). Normalized wastewater data (RSV GCs/1,000 persons/24 h) was correlated to various RSV-incidence timepoints before and after sample collection for all WWTPs. The correlation between the incidence of RSV in the sampling week and RSV in wastewater varied from 0.412 (p = 0.074) to 0.865 (p < 0.001) between the WWTPs. In one study, the Kendall’s τ coefficient for the statewide clinical positivity rate of RSV and RSV B concentrations in wastewater was 0.57 (p < 0.001) (Boehm et al., 2022). Another study reported the Kendall’s τ coefficient between the statewide clinical positivity rate of RSV and RSV concentrations in wastewater to be 0.77 (p < 0.001) for one WWTP and 0.65 (p < 0.001) for another WWTP (Hughes et al., 2022). To continue, Ahmed et al. (2023) showed the Spearman rank correlation between statewide RSV cases and RSV wastewater data to be between 0.39 and 0.95 for the individual WWTPs and 0.53 for the pooled data. In addition, Ando et al. (2023) found the Spearman rank correlation between RSV cases in the city of a given WWTP and the RSV wastewater data to be 0.36–0.52. Thus, the correlations between the clinical data and wastewater data in this study are similar to previously reported values, although one study did not find a statistical correlation between RSV wastewater data and cases. However, the data were correlated with bronchiolitis cases, which is not a specific indicator of RSV cases (Toribio-Avedillo et al., 2023). Furthermore, Hughes et al. (2022), Ahmed et al. (2023), and Ando et al. (2023) show a phenomenon similar to that revealed in this study; there are differences in the correlation coefficients between different WWTPs.

**Table 2.**
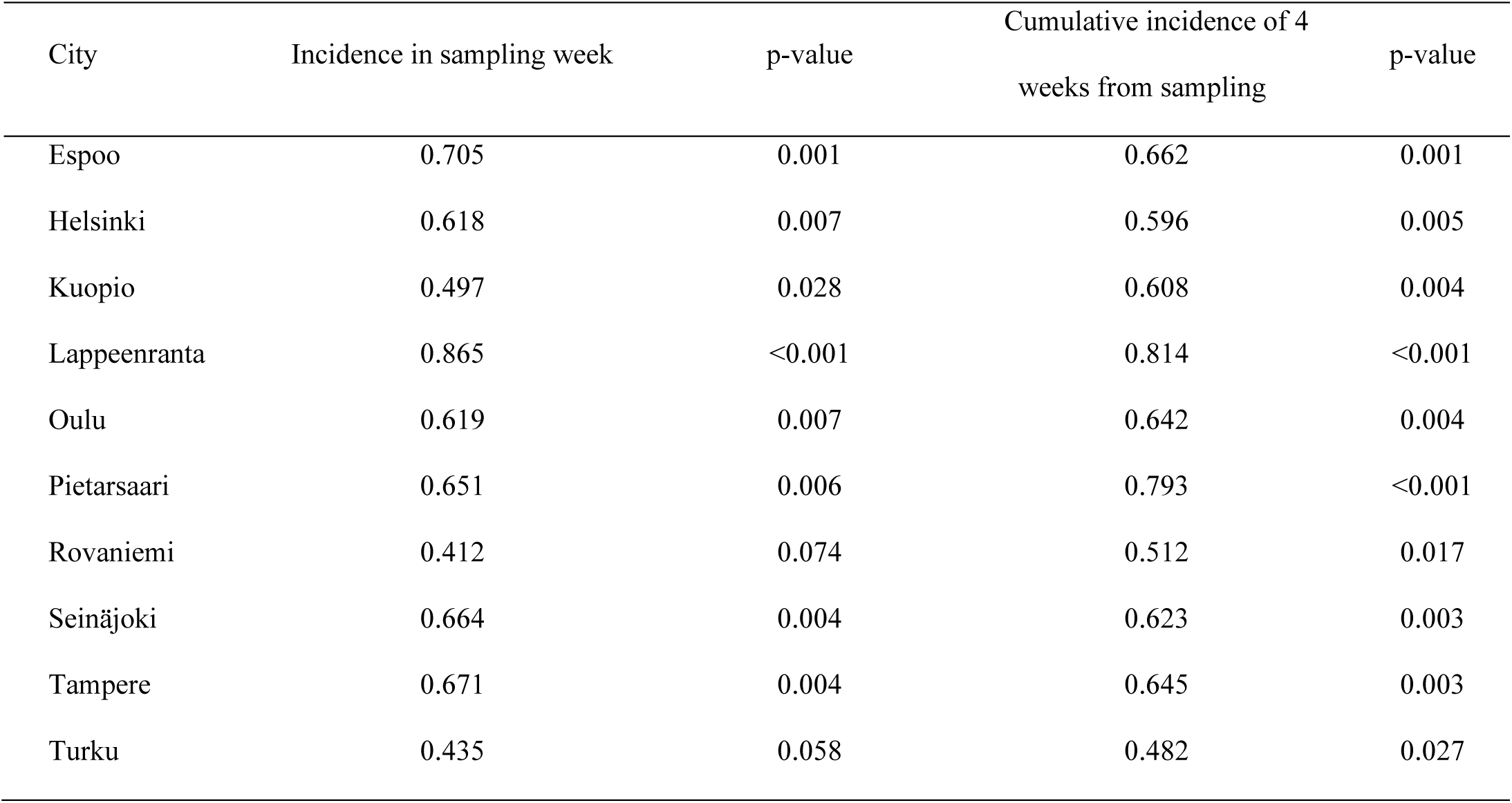
RSV in wastewater (GCs/1000persons/24h) was correlated to the incidence of RSV (RSV cases/100 000 persons/week) and to the cumulative incidence of RSV (RSV cases/100 000 persons/4 weeks from sampling). Normalized wastewater data had a positive correlation to the incidence of RSV. In 50% of the cities, the wastewater data correlated the best with the incidence of the sampling week. In 50% of the cities, the wastewater data correlated better with the cumulative incidence.

In addition to correlating the wastewater results to the incidence on the sampling week, the wastewater data were also correlated with the cumulative incidence for the 4 weeks after the sampling timepoint. The correlation coefficients were similar to those from the sampling week (Table 2). In 50% of cases, the correlation with the cumulative incidence was higher than that with the sampling week. This highlights the capability of wastewater to estimate upcoming cases and the course of an epidemic.

The correlation between viral RNA in wastewater and disease cases, as well as the capability of WBS to predict cases, have also been shown with other viruses, in addition to RSV. Multiple studies have shown that the amount of SARS-CoV-2 RNA in wastewater is correlated with the number of COVID-19 disease cases (Acosta et al., 2023; Tanimoto et al., 2022; Wani et al., 2023; Wolfe et al., 2021; Zhan et al., 2022). The correlation between wastewater and clinical cases has also been found for the influenza virus (Boehm et al., 2022; Wolfe et al., 2022; Wolken et al., 2023). In Finland, the correlation coefficient of influenza A RNA in wastewater and the incidence of influenza A (CC = 0.427–0.870) was shown to be similar to the RSV correlation measured in our study (CC = 0.412–0.865) (Lehto et al. 2023). The study was conducted using the same WWTPs as this study. Similarly, in the influenza A study, the correlation between wastewater data and clinical data varied between WWTPs.

### 3.5 Limitations of the study

The main limitation of the study is the low frequency of the sample collection. The resolution of monthly wastewater data is not optimal as compared to the daily NIDR data. Monthly sampling does not allow for the accurate monitoring of the initiation, peak and end of an epidemic precisely. In 50% of the WWTPs, we could not detect the RSV epidemic from wastewater before the incidence of RSV exceeded five cases/100,000 persons/week. Still, RSV was detected in wastewater in all the cities before the epidemic reached its peak. Furthermore, as the wastewater data correlated with the cumulative incidence during the 4 weeks after sampling, wastewater could potentially be used to estimate the development of epidemics, and it cannot be ruled out that, with more regular sampling, it could be possible to detect the beginning of an epidemic from wastewater early on. This is also supported by other studies (Ahmed et al., 2023; Boehm et al., 2022; Hughes et al., 2022).

One other potential limitation is the sample material, the supernatant fraction of the wastewater. Some previous RSV studies and many other viral studies of wastewater have used settled solids as the sample material (Boehm et al., 2022; Hughes et al., 2022). Our study was able to detect RSV from the supernatant successfully, showing that the supernatant can be used.

One other limitation of this study is the underestimation of the RSV cases in the NIDR. RSV is a respiratory infectious disease, and many cases are asymptomatic or have only mild symptoms. Therefore, all cases are not diagnosed in healthcare. Furthermore, the symptoms of RSV are similar to those of other respiratory viruses, and RSV-specific laboratory tests are needed for the diagnosis of the disease. These factors may lead to inaccuracies, for example, in the temporal comparison of virus GC numbers in wastewater and NIDR data. Still, our study used the RSV cases of the WWTP catchment area, not the city or the state of the WWTP. This provides more accuracy in the estimation of RSV incidence as compared to previous studies.

### 3.6 The significance of using WBS in the surveillance of RSV

RSV is the most common virus that causes respiratory infections in young children. The elderly and immunocompromised people are also at-risk groups. Due to the COVID-19 pandemic, there was no RSV epidemic in 2020–2021, which resulted in a large epidemic in 2021–2022. The burden of RSV on children’s healthcare in Finland was even higher than that of COVID-19, and it was the most common reason for the hospitalization of children during the 2021–2022 RSV epidemic (Helsinki University Hospital, 2022). In this study, WBS was shown to be a useful tool for use in tracking the RSV epidemic at the national level in Finland. The WBS of RSV was shown to reflect the number of RSV cases, and it has the potential to be used as an early-warnings tool. However, weekly sampling would be needed for the more precise monitoring of the disease. WBS is recognized as a useful public health monitoring tool for SARS-CoV-2 (Agrawal et al., 2021; Keshaviah et al., 2021). Similarly, our study shows that the wastewater monitoring of RSV provides useful information for public health monitoring and helps authorities in decision-making.

The effective use of WBS for viruses requires efficient information sharing, for example, through national sentinel systems. The European commission has already advised that wastewater should be monitored for SARS-CoV-2, poliovirus, and influenza virus (EU, 2022). In addition to RSV, more information on the capabilities of WBS can be gathered by studying other viruses. Wastewater is known to include multiple disease markers, not only for communicable diseases but also for noncommunicable diseases. Maybe, in the future, even noncommunicable diseases could be studied from wastewater.

## Author Contributions

The manuscript was written based on the contributions of all authors. All authors have approved the final version of the manuscript.

Annika Länsivaara: conceptualization, methodology, formal analysis, investigation, data curation, writing–original draft and editing; Kirsi-Maarit Lehto: conceptualization, methodology, investigation, data curation, writing–original draft, review, and editing, supervision, project administration, funding acquisition; Rafiqul Hyder: methodology, investigation, writing– original draft, review, and editing; Oskari Luomala: Investigation, Formal analysis, writing– review and editing; Anssi Lipponen: writing–review and editing, project administration; Annamari Heikinheimo: writing–review and editing, project administration, funding acquisition; Tarja Pitkänen: writing–review and editing, project administration, funding acquisition; Sami Oikarinen: conceptualization, methodology, formal analysis, investigation, data curation, writing–original draft, review and editing, supervision, project administration, funding acquisition.

## Funding Sources

This work was funded by Academy of Finland, grant number 339416.

## Data Availability

All data produced in the present study are available upon reasonable request to the authors

## Acknowledgements

The authors thank Annika Laaksonen for her assistance with the laboratory work.

The authors would like to express special thanks to the personnel of the WWTPs included in the WastPan study.

## WastPan study group

Tampere University: Sami Oikarinen, Kirsi-Maarit Lehto, Rafiqul Hyder, Annika Länsivaara, and Erja Janhonen.

The Finnish Institute for Health and Welfare: Tarja Pitkänen, Ananda Tiwari, Anssi Lipponen, Anna-Maria Hokajärvi, Anniina Sarekoski, Aleksi Kolehmainen, Teemu Möttönen, Oskari Luomala, Aapo Juutinen, Soile Blomqvist, Kati Räisänen, and Carita Savolainen-Kopra.

University of Helsinki: Annamari Heikinheimo, Ahmad Al-Mustapha, Viivi Heljanko, Venla Johansson, and Paula Kurittu.

### Abbreviations

RSV: Respiratory syncytial virus
NIDR: National Infectious Diseases Register
WWTP: Wastewater treatment plant
WBS: Wastewater-based surveillance
RT-qPCR: Reverse-transcription quantitative polymerase chain reaction

## Notes

### Competing Interest Statement

Sami Oikarinen reports a relationship with Greenseq Ltd that includes: board membership. Kirsi-Maarit Lehto reports a relationship with Greenseq Ltd that includes: board membership and travel reimbursement.

### Author Declarations

The clinical RSV data was retrieved from the Finnish National Infectious Diseases register: https://sampo.thl.fi/pivot/prod/fi/ttr/shp/fact_shp. The Finnish Institute for Health and Welfare took part in this study and granted permission to use the data.

